# Dual tasking exacerbates force and neural control unsteadiness in sarcopenic older adults

**DOI:** 10.64898/2026.04.28.26350825

**Authors:** Lucas B. R. Orssatto, Brian C. Clark, David Scott, Hélio V. Cabral, Gabriel L. Fernandes, Robin M. Daly

## Abstract

**Background:** Sarcopenia is associated with impaired physical function. Dual-task conditions, which increase cognitive demand during motor performance, may reveal deficits in neuromuscular control that are not evident during isolated motor tasks. Therefore, we investigated whether older adults with sarcopenia exhibit poorer steadiness of force and neural control (i.e., greater motor unit discharge variability, and altered common synaptic input) during submaximal contractions performed under single- and dual-task conditions compared with non-sarcopenic controls and master athletes.

**Methods:** Fifty-two older adults were included (74.3±7.3 years; 50% female). Sarcopenia was defined using Sarcopenia Definitions and Outcomes Consortium criteria based on low grip strength and slow gait speed. Participants (11 with sarcopenia, 22 controls, and 19 masters athletes) performed six sustained isometric ankle dorsiflexion contractions at 30% maximal voluntary torque, three under single-task conditions and three during concurrent serial number subtraction. High-density surface electromyography was recorded from tibialis anterior, and motor unit spike trains were decomposed and tracked across trials. Outcomes included torque coefficient of variation, mean discharge rate, inter-spike interval coefficient of variation, and intramuscular coherence in the delta (1–5 Hz), alpha (5–15 Hz), and beta (15–35 Hz) bands.

**Results:** Sarcopenic individuals had worse torque steadiness (increased torque coefficient of variation) than controls (45-84%) and athletes (39-105%) during single-task, which worsened further (+35% relative to baseline) during dual-tasking. Mean discharge rates (proxy of neural drive) slightly increased during dual-tasking in all groups by ∼2.6%, with no between-group differences. Discharge rates coefficient of variation (Proxy of neural control unsteadiness) increased 5.5% in sarcopenia, was unchanged in controls, and decreased 4.1% in athletes during dual-tasking. Delta-band coherence decreased 5.5% during dual-tasking across all groups. Alpha-band coherence increased only in sarcopenia during dual-tasking (20.6%). Beta-band coherence increased 20.6% in sarcopenia but decreased 3.6% in controls and 3.8% in athletes during dual-tasking.

**Conclusions:** Older adults with sarcopenia exhibit poorer force and neural control steadiness, and both deficits worsen under cognitive load. These changes are accompanied by alterations in common synaptic input, particularly an increase in physiological involuntary tremor-related oscillations (alpha band), which contribute to greater force unsteadiness. Neural control unsteadiness during dual-task performance may therefore represent a neural feature of sarcopenia-related functional impairment. Assessing neuromuscular control during cognitively demanding tasks may improve detection of neural dysfunction and identify mechanistic targets for interventions to reduce mobility impairment and fall risk. These findings support expanding muscle-centric views of sarcopenia to include neural mechanisms of motor control.

## INTRODUCTION

Sarcopenia is a progressive skeletal muscle disease that has been strongly linked to frailty, falls, disability, and loss of independence in older adults^1^. Although it is commonly defined by low muscle mass, low muscle strength, and impaired physical function^1–3^, declines in force-producing capacity and control are unlikely to be explained by muscle morphology alone^4–6^. Deterioration in the neuromotor system^4,5,7–9^ is functionally relevant because stable force production depends on the nervous system’s ability to generate and maintain coordinated neural drive to muscle^10^. Consistent with this, older adults exhibit greater force fluctuations and more variable motor unit discharge during submaximal sustained muscle contraction than young adults, suggesting that impaired neural control is a key feature of age-related reductions in motor performance^7,11–14^. These deficits may be especially important under conditions that more closely reflect everyday function or activities of daily living

Many real-world activities relevant to falls and independence, such as walking, rising from a chair, or carrying objects, are performed while attention is divided between motor and cognitive demands^11,12^. Under such dual-task conditions, older adults commonly move more slowly, show greater instability, and are more likely to make contact with obstacles^15–18^, with impaired dual-task performance also linked to both prior falls and future fall risk^15,19^. Experimental work further shows that adding a cognitive task during steady contractions increases force and motor unit discharge variability, indicating that cognitive load can degrade the neural control of force^11,12^. Individuals with sarcopenia also demonstrate poorer dual-task gait performance than their non-sarcopenic peers^18^. In contrast, older adults engaged in long-term sports training (i.e., masters athletes), which typically involve multi-tasking while performing, exhibit better resilience to dual-task interference^20,21^. We have previously shown that when comparing heterogeneous phenotypes of older adults (i.e., sarcopenic, non-sarcopenic controls, and masters athletes) there are important contributions (differences) in the intrinsic motoneuron excitability to poor physical function^22^. However, whether these phenotypic differences in physical function are also accompanied with altered and force control, particularly under dual-task conditions, is not known.

High-density surface electromyography (HD-sEMG) combined with motor unit decomposition enables detailed examination of motor unit discharge behaviour during voluntary contractions^23^. By integrating measures of torque steadiness and motor unit discharge properties, it is possible to quantify both the steadiness of motor output and the organisation of common synaptic inputs to the motoneuron pool^10,11^. Common synaptic input refers to neural signals that are sent to many motor units, particularly from descending corticospinal pathways^24,25^, such that their resultant activity is highly correlated^26,27^. This shared input therefore determines the effective neural command to the muscle, enabling smooth and coordinated force production^10,28^. Estimating the degree of shared input provides complementary mechanistic information into how neural commands to the muscle are organised over time, and how its disruption generate greater fluctuations in motor output and poorer force control, reflecting the combined influence of neural pathways at different levels of the nervous system^10^.This approach may therefore help determine not only whether neural control is impaired in those with sarcopenia, but also whether cognitive loading reveals abnormalities in the organisation of neural drive that are not evident under single-task conditions. Thus, the aim of this study was to characterise neuromuscular control during submaximal isometric contractions performed under single- and dual-task conditions in older adults with sarcopenia, non-sarcopenic controls, and master athletes. We hypothesised that participants with sarcopenia would exhibit poorer torque steadiness and greater motor unit discharge variability, particularly under cognitive load, whereas master athletes would demonstrate the most stable performance. We further hypothesised that dual-tasking would alter common synaptic input in sarcopenia, consistent with a reduced capacity to maintain coordinated neural drive when cognitive demands are increased.

## METHODS

### Participants

This cross-sectional study was conducted in the same cohort described previously^22^, but used a distinct dual-task isometric protocol and motor unit analysis to address a separate mechanistic question. Full participant characterisation is reported elsewhere^22^. Eligibility criteria included age ≥65 years, no use of medications affecting noradrenergic or serotonergic systems (e.g., psychiatric medications), no sex hormone replacement therapy or performance-enhancing drugs, and no lower-limb musculoskeletal or neurological conditions that could affect testing outcomes. Sarcopenic individuals and controls required a body mass index ≤30 kg/m^2^. Master athletes had ≥10 years of competitive sport participation (with <1 year interruption) and engagement in competition within the previous year. Controls were non-sarcopenic and reported ≤1 structured exercise session per week in the previous year. The study was approved by the Deakin University Human Research Ethics Committee (2023–241) and conducted in accordance with the Declaration of Helsinki. All participants provided written informed consent.

### Sarcopenia screening

Participants underwent a multi-stage screening protocol to determine sarcopenia status^22^. Initial screening used the five-item SARC-F questionnaire^29^. Individuals scoring ≥2 completed additional questions on walking ability and load carrying, and a video-based Five Times Sit-to-Stand (5STS) test^30^. Those reporting slow walking, difficulty lifting small loads, and a 5STS time >15 s were invited for in-person assessment. All participants underwent the laboratory screening workflow, allowing the sarcopenia group to be classified and confirming that all control and masters groups participants were non-sarcopenic according to SDOC criteria^2^. Sarcopenia was diagnosed according to Sarcopenia Definitions and Outcomes Consortium (SDOC) criteria^2^, defined by low muscle strength (handgrip strength <20 kg for females or <35.5 kg for males) and impaired physical performance (usual gait speed <0.8 m·s□^1^). Handgrip strength was measured with a digital dynamometer (Jamar Plus, USA) with participants seated, shoulder neutral and elbow flexed at 90°. Participants performed two maximal contractions (3–5 s) with the dominant hand, and the highest value was used. Handgrip strength (kg) was measured with a digital dynamometer (Jamar Plus, USA) with participants seated, shoulder neutral and elbow flexed at 90°. Participants performed two maximal contractions (3–5 s) with the dominant hand, and the highest value was used.

### Neuromuscular assessments

Participants were seated upright with their preferred leg secured to an ankle dynamometer (DinamometroGC, OT Bioelettronica, Italy) with the hip at ∼70° flexion, knee at 0° extension, and ankle at 5° plantarflexion (0° anatomical position). The foot was strapped below the proximal phalanx and the knee secured to prevent movement. Skin over the tibialis anterior was shaved, abraded (Everi preparation paste, Spes Medica, Italy), and cleaned with 70% isopropyl alcohol. Two HD-EMG 64-channel electrode grids (8-mm interelectrode distance; HD08MM1106, OT Bioelettronica) were placed over the superior and inferior regions of the tibialis anterior using bi-adhesive foam and conductive paste (AC cream, Spes Medica). A compression bandage secured electrode contact and a dampened strap electrode (WS1.1, OT Bioelettronica) placed around the ankle served as reference.

Data collection began after a 10–15 min rest following a previous experiment that tested maximal and submaximal triangular-shaped contractions^22^. Participants were familiarised with trapezoidal dorsiflexion contractions (3-s ramp up, 10-s plateau, 3-s ramp down) at 30% of maximal voluntary torque (MVT) determined previously^22^. They were also familiarised with the cognitive task, in which they were asked to subtract 7, continuously, starting from a number between 70 and 99 (except the multiples of 7). All participants were able to perform the subtraction task taking less than 3s to provide an answer after each subtraction. After 5 min rest, participants performed six contractions at 30% MVT (3-s ramp up, 20-s plateau, 3-s ramp down; 2-min rest), three under single-task (motor task alone) and three under dual-task (Motor + cognitive task) conditions in alternating randomised order (Figure 1). Trials were considered invalid if subtraction was interrupted for >3 s; errors of ±1 were accepted, whereas more than one error of ±2 invalidated the trial. One additional trial per condition was allowed to avoid fatigue. During these contractions, high-density surface electromyography was recorded in monopolar mode, amplified (205×), band-pass filtered (10–500 Hz), and digitised at 2000 Hz using a 16-bit wireless amplifier (Muovi+, OT Bioelettronica) and OTBioLab+ software (v1.5.9.2) with real-time visual torque feedback.

**Figure 1.**
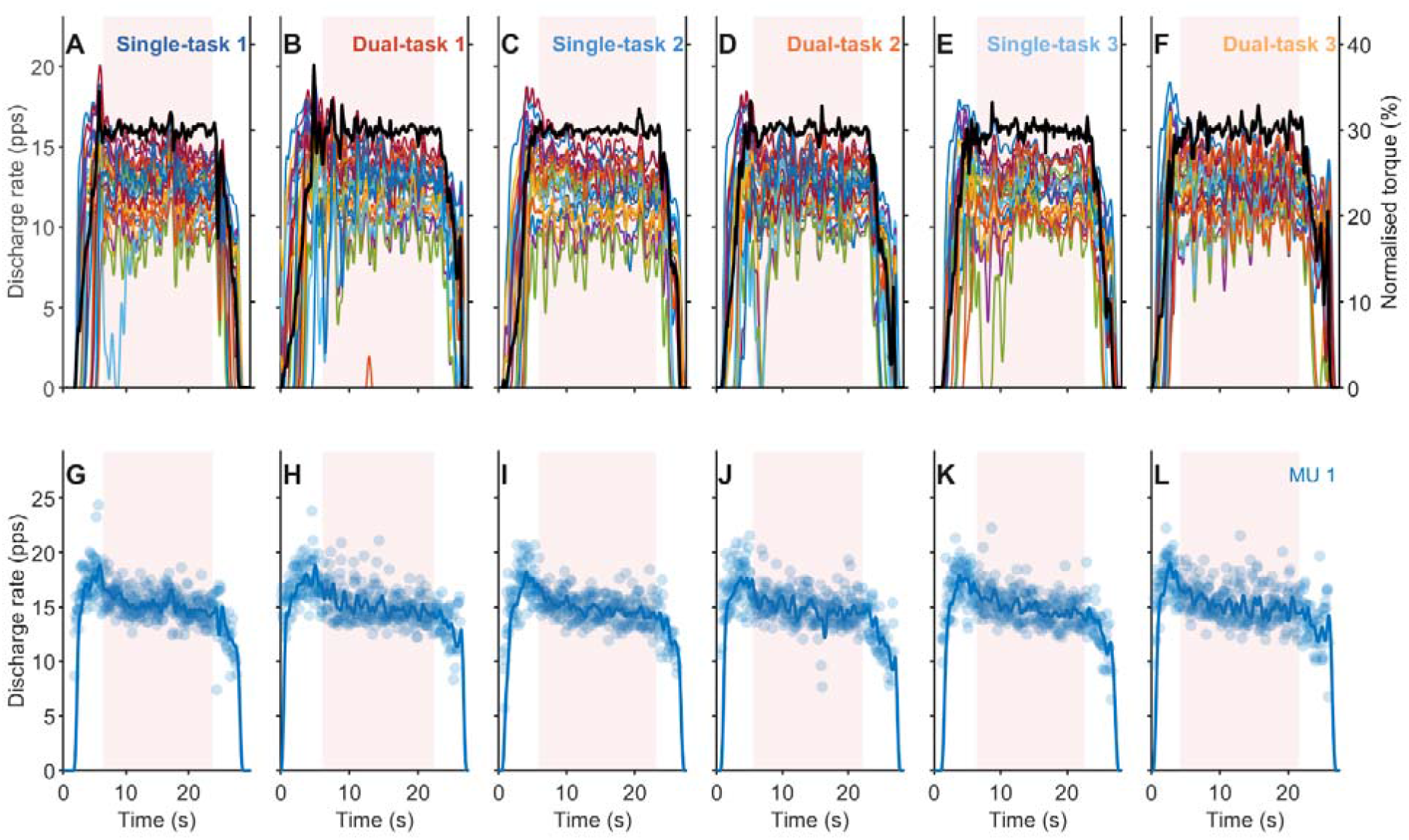
Study design and individual sarcopenic participant data. (A–F) Smoothed discharge rates of individual motor units (coloured lines) identified from the tibialis anterior during isometric ankle dorsiflexion at 30% of maximal voluntary torque. Each panel corresponds to one trapezoidal contraction, with single-task (A, C, E) and dual-task (B, D, F) conditions displayed separately. The black line represents the torque trace (right y-axis), expressed as a percentage of maximal voluntary torque. The shaded red region denotes the analysis window selected at the torque plateau. (G–L) Instantaneous discharge rate (dots) and smoothed discharge rate (solid line) of the earliest-recruited motor unit across the six contractions, corresponding to panels A–F respectively. The shaded red region indicates the analysis window used for coherence estimation. Units that did not exhibit continuous firing throughout the analysis window (e.g., light blue unit from panel A and orange unit from panel B) were excluded from statistical analyses but are still displayed in panels A–F for transparency and to illustrate the full pool of identified motor units.

### Torque and HD−EMG analyses

The load cell signal from the ankle dynamometer was digitised, amplified (205×), and low-pass filtered offline at 15 Hz. HD-EMG signals were processed offline using DEMUSE software^31^ and band-pass filtered (20–500 Hz) with a second-order zero-lag Butterworth filter. Motor unit spike trains were identified using the convolutive kernel compensation algorithm^31^. Separation vectors were then applied to concatenated HD-EMG signals to track motor units across trials and conditions^32^. Duplicate units (>30% common discharge times) were removed^33^. An experienced investigator (GLF), blinded to group and condition, manually corrected missed or misidentified spikes. Only motor units with a pulse-to-noise ratio ≥30 dB were included. Units with non-continuous firing during the analysis window were not included in the analyses.

Instantaneous discharge rates were calculated as the inverse of inter-spike intervals. Variables were extracted from an ∼18 s window during the plateau phase of the contraction (excluding the first and last seconds): (i) torque coefficient of variation (%), (ii) mean discharge rate, (iii) inter-spike interval coefficient of variation (%), and (iv) intramuscular coherence metrics (see below). Motor unit recruitment threshold was defined as the torque (%MVT) at the first discharge.

Intramuscular coherence between motor unit spike trains was used to estimate common synaptic oscillations to the motoneuron pool^34,35^. Coherence was computed between two cumulative spike trains (CSTs) formed by summing the binary discharge trains of randomly selected motor units. For each participant, each CST comprised five motor units^36^, and the procedure was repeated for up to 100 random permutations to obtain pooled estimates. For each permutation, coherence was calculated between detrended CSTs using Welch’s periodogram with a 1 s Hanning window and 95% overlap^35^. Coherence spectra were averaged across permutations and transformed into z-scores^37^. Only z-scores exceeding the bias threshold were retained, defined as the mean z-coherence between 250–500 Hz, a frequency range with no expected physiological coherence^35,36^. Differences in common synaptic input between conditions were quantified as the area under curve (AUC) of the z-coherence within the delta (δ, 1–5 Hz), alpha (α, 5–15 Hz), and beta (β, 15–35 Hz) bands (Figure 2). Low-frequency delta-band coherence reflects common drive associated with force fluctuations^10^ alpha-band activity has been linked to subcortical and spinal contributions related to involuntary physiological tremor; and beta-band coherence is commonly associated with corticospinal coupling^10^. These bands are the frequency ranges most consistently examined in motor unit coherence studies and have been linked to different temporal features of common neural input to the motoneuron pool^10,28,35^. However, because coherence does not permit definitive source localisation, these bands were interpreted conservatively as indexing frequency-specific organisation of common input rather than discrete anatomical generators.

**Figure 2.**
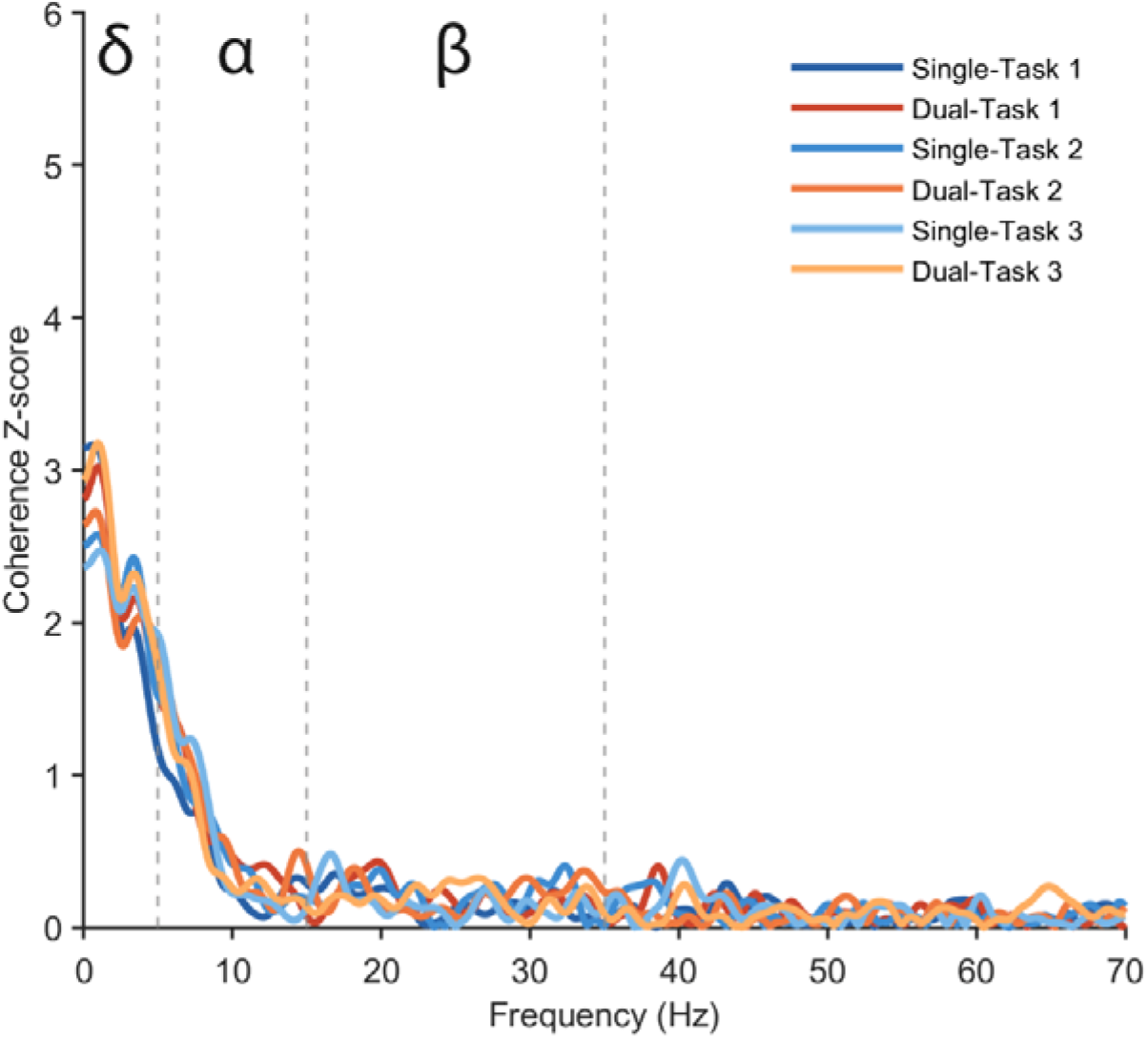
Coherence Z-scores during single-task and dual-task isometric ankle dorsiflexion in a participant with sarcopenia. Coherence Z-scores as a function of frequency (0–70 Hz) computed from the cumulative spike trains of motor units identified from the tibialis anterior during isometric contractions at 30% of maximal voluntary torque. Each line represents one trapezoidal contraction, with single-task trials shown in blue shades (Single-Task 1–3) and dual-task trials shown in orange-red shades (Dual-Task 1–3). Dashed vertical lines delineate frequency bands of interest: delta (δ, 1–5 Hz), alpha (α, 5–15 Hz), and beta (β, 15–35 Hz).

### Statistical analysis

All analyses were conducted using robust linear mixed-effects models to mitigate violations of classical linear model assumptions and downweigh the effects of outliers. Robust estimates were obtained via M-estimation within linear mixed-effects models. Primary inference emphasised point estimates and 95% confidence intervals (CIs) rather than null-hypothesis significance testing.

Separate robust linear mixed−effects models for force and motor unit-derived variables tested group (athletes, controls, and sarcopenia) by condition (Single and dual tasks) effects and their interaction, adjusting for sex (fixed effect) and motor unit recruitment threshold (covariate) when appropriate. Each trial and motor units (for mean discharge rates and inter-spike interval coefficient of variation) were nested within participants, and random intercepts were included to account for intra-individual correlation. For each contrast, bootstrap CIs were computed with 2,000 resamples. Interpretation focused on the magnitude and precision of mean differences; effects were considered noteworthy when CIs excluded zero. The effect of age as a covariate was tested on each model. Since it did not change the results for any outcome, age was not included in the final models. The results of the models accounting for age are reported in Supplementary Material 1. An exploratory analysis was conducted comparing subgroups of endurance- and power-type athletes, reported in Supplementary Material 2.

All analyses and figure generation were conducted using RStudio (version 2024.04.2) and MATLAB (Version 2023b, The MathWorks, Natick, MA, USA). The dataset, R code, and packages information can be found at: https://github.com/orssatto/mucoh-sarcopenia.

## RESULTS

### Participants and motor unit yield

Fifty-six participants were initially tested. Four were excluded (one sarcopenic and one control unable to perform the dual task; two athletes withdrew due to time constraints). Eleven sarcopenic (mean±standard deviation age, 80.1±8.1 years, and body mass, 64.6±13.1kg), 22 controls (74.8±6.5 years, 70.7±13.7kg), and 19 master athletes (70.3±6.1 years, 69.7±15.0kg) were included in the analysis.

A total of 8,302 motor unit spike trains were identified across contractions and conditions, derived from 251 motor units in sarcopenic participants, 638 in controls, and 735 in athletes. The median (1st–3rd quartiles) number of motor units per participant was: sarcopenic—females 18 (11–24), males 31 (27–34); controls— females 22 (20–27), males 38 (31–44); athletes—females 27 (19–32), males 46 (41–50).

### Torque and high-density electromyography outcomes

#### Torque steadiness was reduced in sarcopenia and worsens during dual tasking

Force control was less steady (i.e., higher torque coefficient of variation) in the sarcopenic group than in controls and athletes during both single- and dual-task conditions (Figure 3). Force steadiness worsened during the dual-task condition in the sarcopenic group only, whereas no condition differences were observed in controls or athletes. Figure 3 shows estimated marginal means and mean differences in torque coefficient of variation (a proxy of force steadiness) for each group and condition. The statistical model identified a group by condition interaction [β = -1.37 (-1.84, -0.91), t = -5.75], with no effect of sex [β = 0.06 (-0.524, 0.646), t = 0.20].

**Figure 3.**
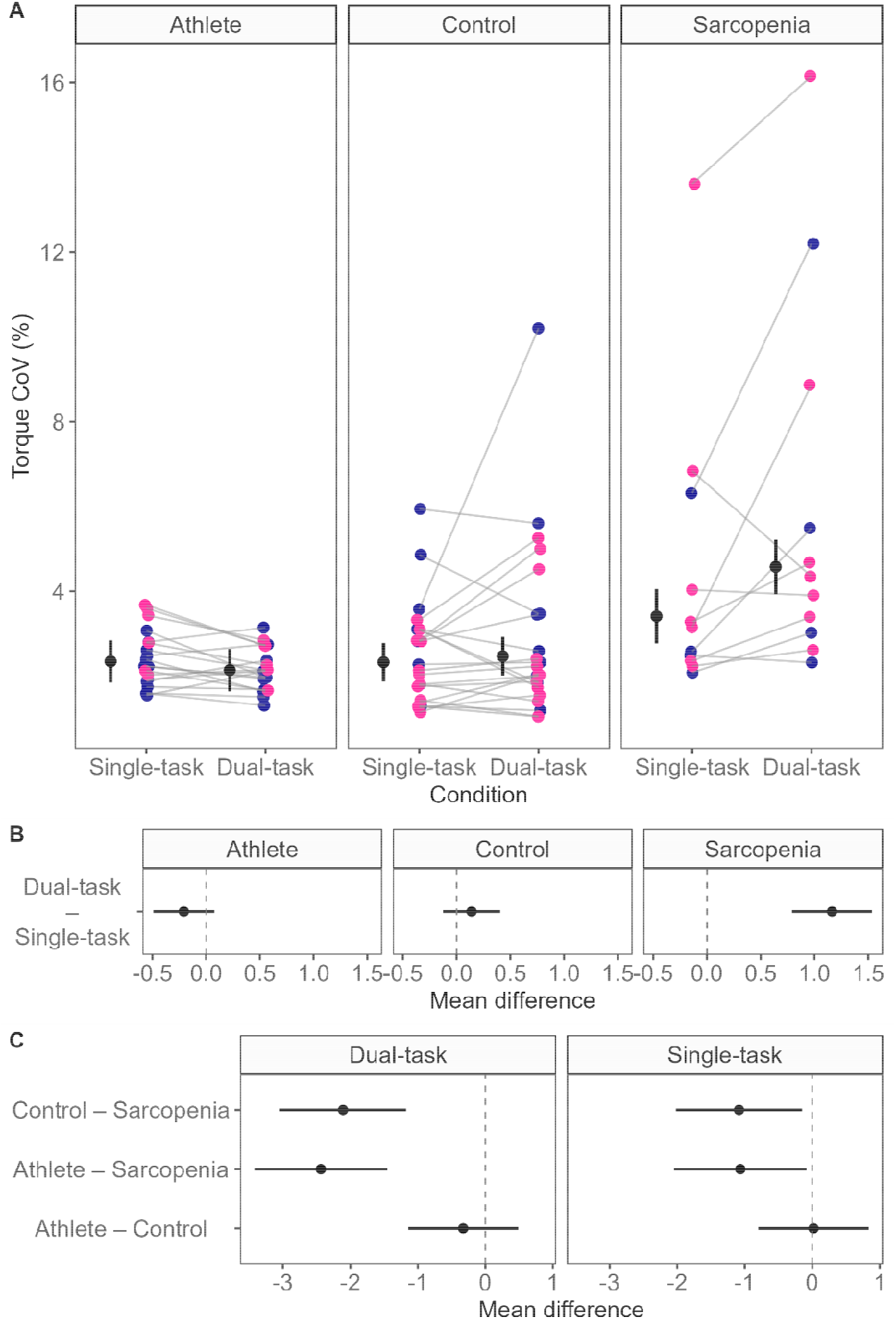
Effects of dual-tasking on torque coefficient of variation (CoV) across groups. Panel A displays participant-level torque CoV during single-task and dual-task conditions for Athlete, Control, and Sarcopenia groups. Each coloured point represents the mean discharge rate of one participant (blue = male, pink = female). Grey lines connect the same participant across conditions. Panel B displays condition contrasts (dual-task – single-task) shown separately for each group. Torque CoV remained unchanged for athletes and controls, and increased markedly for sarcopenic individuals under dual-tasking. Panel C displays group contrasts, shown separately for dual- and single-tasks conditions. Under dual-tasking, sarcopenic individuals exhibited significantly higher torque CoV compared with both athletes and controls. For all panels, black circles and error bars indicate estimated marginal means ± 95% confidence intervals.

#### Mean discharge rates increased in all groups during dual tasking

Mean discharge rates, a proxy of the neural drive to the muscle, increased during the dual-task condition in all groups, with no differences observed between sarcopenic, control, and athlete participants in either condition. Figure 4 shows estimated marginal means and mean differences in mean discharge rate for each group and condition. The statistical model identified a group by condition interaction [β = 0.27 (0.20, 0.33), t = 8.16]. Effects of recruitment threshold [β = -0.031 (-0.034, -0.028), t = -20.0] and sex were also observed [β = -1.3 (-2.4, -0.3), t = -2.51], indicating that motor units recruited at higher thresholds discharged at lower rates, and that males displayed lower mean discharge rates than females.

**Figure 4.**
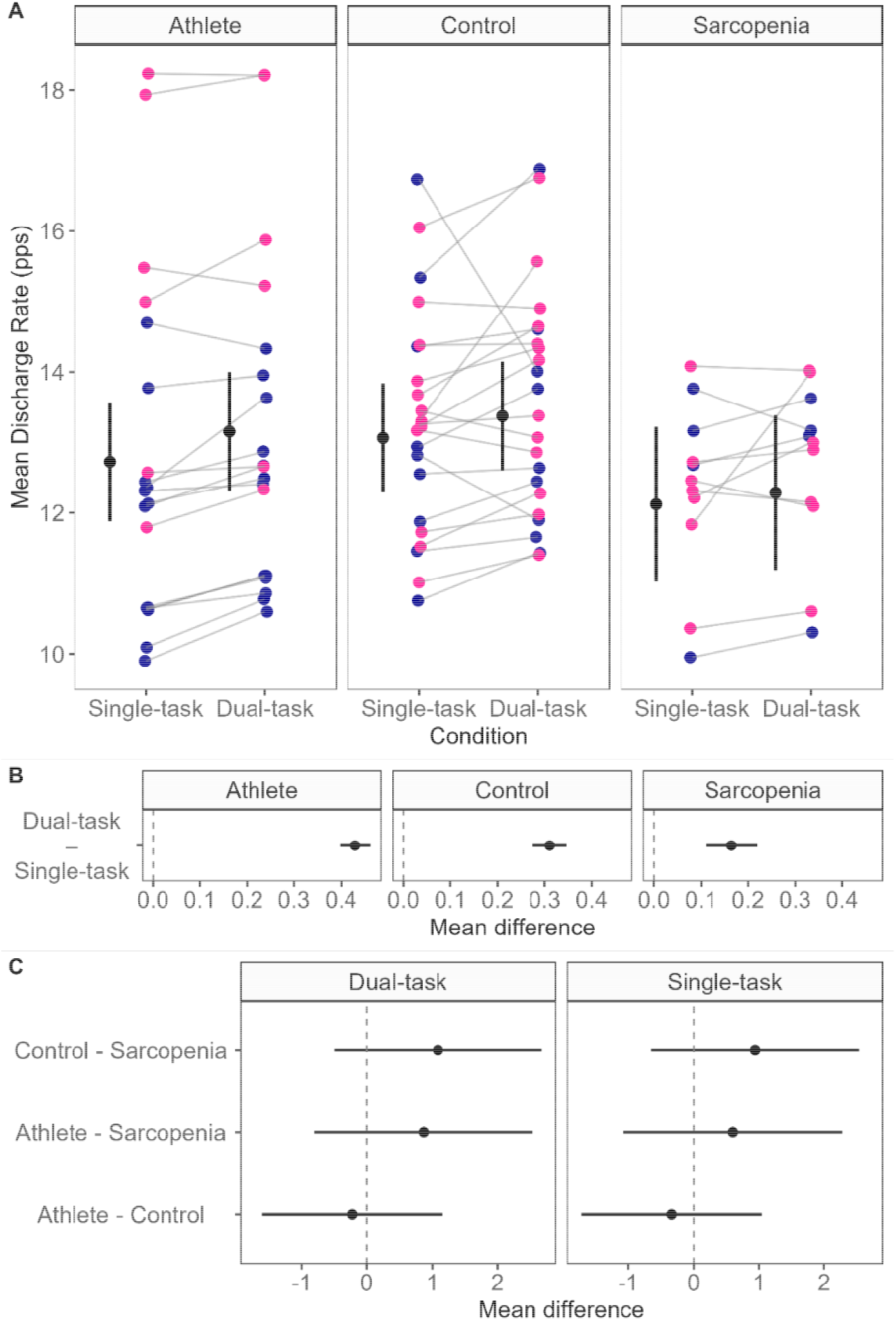
Effects of dual-tasking on mean motor unit discharge rates across groups. Panel A displays participant-level mean discharge rates (pps) during single-task and dual-task conditions for Athlete, Control, and Sarcopenia groups. Each coloured point represents the mean discharge rate of one participant (blue = male, pink = female). Grey lines connect the same participant across conditions. Panel B displays condition contrasts (dual-task – single-task) shown separately for each group. Mean discharge rates were significantly increased during dual-tasking compared with single-tasking across all groups. Panel C displays group contrasts, shown separately for dual- and single-task conditions. No significant group differences were detected in either condition. For all

#### Neural control steadiness worsened in sarcopenic individuals, improved in athletes, and remained unchanged in controls during dual-tasking

Neural control was less steady (i.e., higher inter-spike interval coefficient of variation) in the sarcopenic group than in controls and athletes during both single- and dual-task conditions (Figure 5). Neural drive steadiness worsened during the dual-task condition in the sarcopenic group, improved in athletes, and remained unchanged in controls. Athletes displayed more stable neural drive (lower coefficient of variation) than controls during the dual-task condition only. Figure 5 shows estimated marginal means and mean differences in inter-spike interval coefficient of variation (as a proxy of neural drive steadiness) for each group and condition. The statistical model identified a group by condition interaction [β = -1.53 (-1.80, - 1.27), t = -11.3]. An effect of recruitment threshold was observed [β = 0.14 (0.13, 0.15), t = 27], indicating that higher-threshold motor units exhibited greater discharge variability, while no effect of sex was detected [β = 0.02 (-1.51, 1.55), = 0.03].

**Figure 5.**
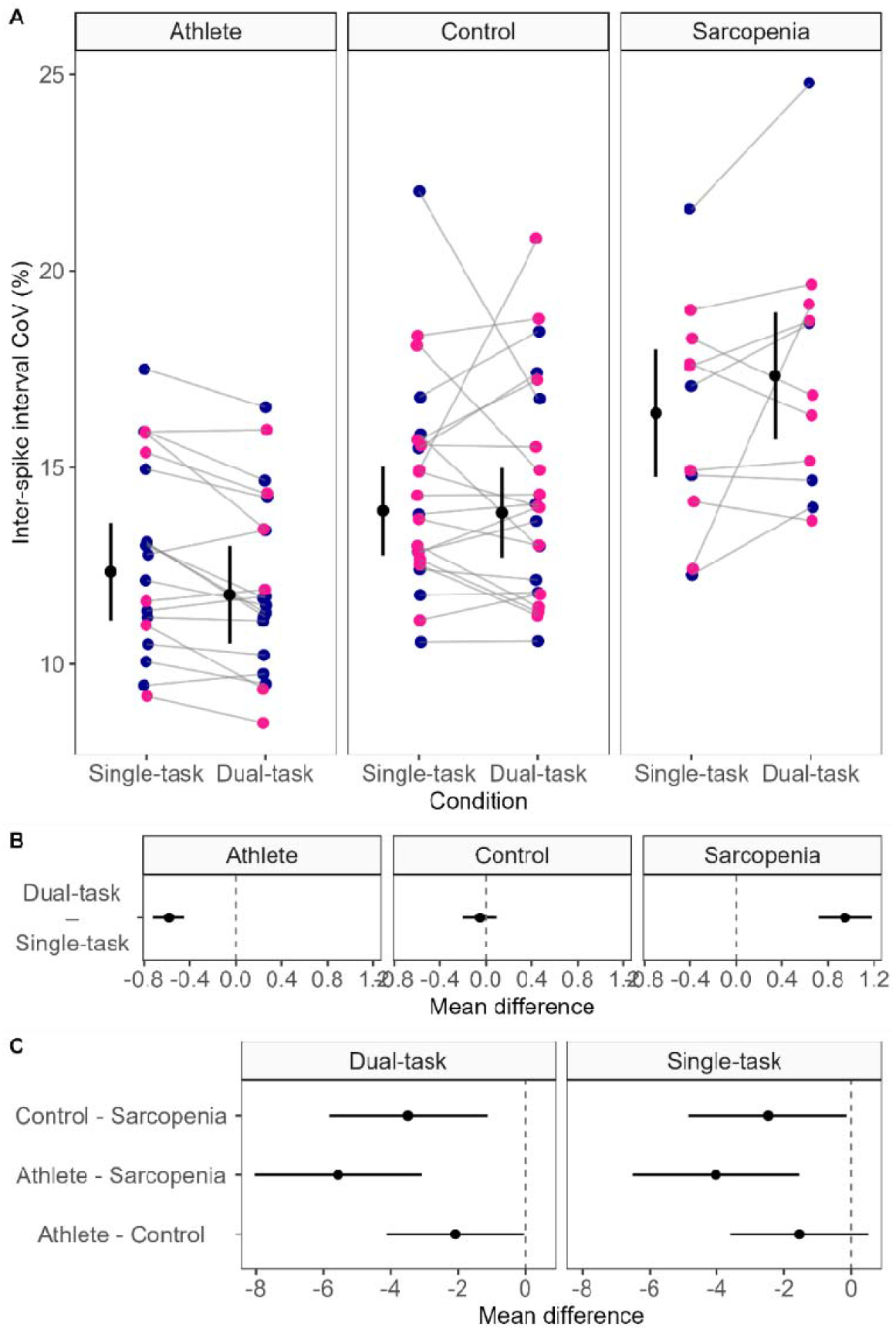
Effects of dual-tasking on inter-spike interval variability across groups. Panel A displays participant-level inter-spike interval coefficient of variation (CoV, %) during single-task and dual-task conditions for Athlete, Control, and Sarcopenia groups. Each coloured point represents the mean value for one participant (blue = male, pink = female). Grey lines connect the same participant across conditions. Panel B displays condition contrasts (dual-task – single-task) shown separately for each group. Inter-spike interval CoV was significantly reduced in athletes, remained unchanged in controls, and increased in sarcopenic individuals under dual-tasking. Panel C displays group contrasts, shown separately for dual- and single-task conditions. Under dual-tasking, sarcopenic individuals exhibited significantly higher inter-spike interval CoV compared with both athletes and controls, whereas athletes demonstrated lower variability than controls. For all panels, black circles and error bars indicate estimated marginal

#### Common input associated with force fluctuations was reduced in all groups during dual-tasking

Common neural input related to force fluctuations (delta-band) was reduced during the dual-task condition in all groups, with no differences observed between sarcopenic, control, and athlete participants in either condition. Figure 6 shows estimated marginal means and mean differences in delta-band coherence for each group and condition. The statistical model identified a condition effect [β = 5.53 (2.31, 8.75), t = 3.37], with no effect of sex [β = -3.04 (-13.9, 7.8), t = -0.55].

**Figure 6.**
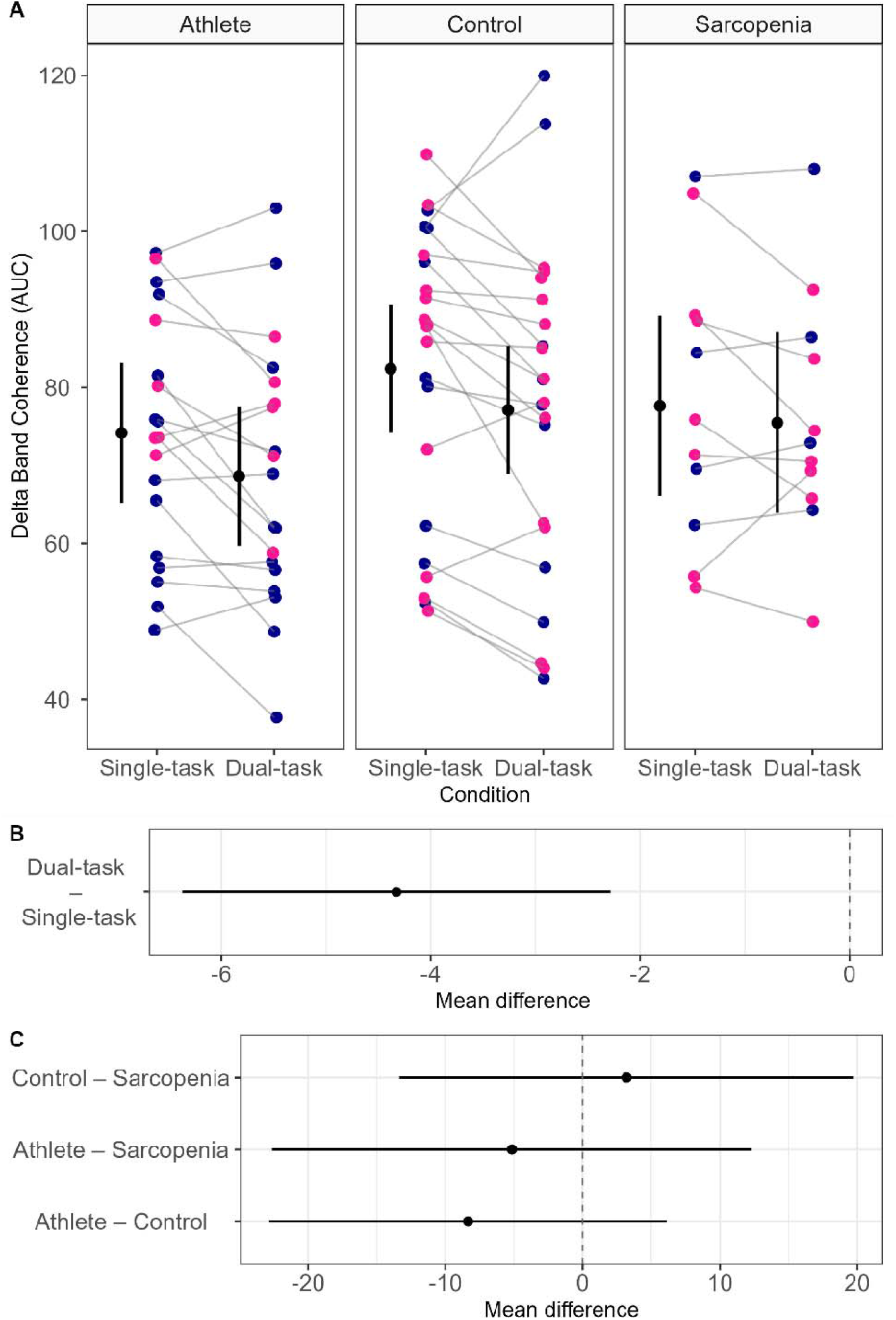
Effects of dual-tasking on delta-band coherence across groups. Panel A displays participant-level delta-band coherence area under the curve (AUC) during single-task and dual-task conditions for Athlete, Control, and Sarcopenia groups. Each coloured point represents the mean value for one participant (blue = male, pink = female). Panel B displays condition contrasts (dual-task – single-task), collapsed across groups. Delta-band coherence was significantly reduced during dual-tasking compared with single-tasking across all groups. Panel C displays group contrasts, collapsed across conditions. No significant differences were observed between groups in either condition. For all panels, black circles and error bars indicate estimated marginal means ±

#### Involuntary physiological tremor fluctuations in common input increased in the sarcopenic group during dual-tasking

Neural input associated to involuntary physiological tremor contributions to common input (alpha-band coherence) increased during the dual-task condition in the sarcopenic group only (Figure 7). No changes were observed between conditions in controls or athletes, and no differences between these groups were detected in either condition. Figure 7 shows estimated marginal means and mean differences in alpha-band coherence for each group and condition. The statistical model identified a group by condition interaction [β = -12.5 (-18.8, -6.2), t = -3.86], with no effect of sex [β = 13.0 (-27.7, 1.7), t = -1.74].

**Figure 7.**
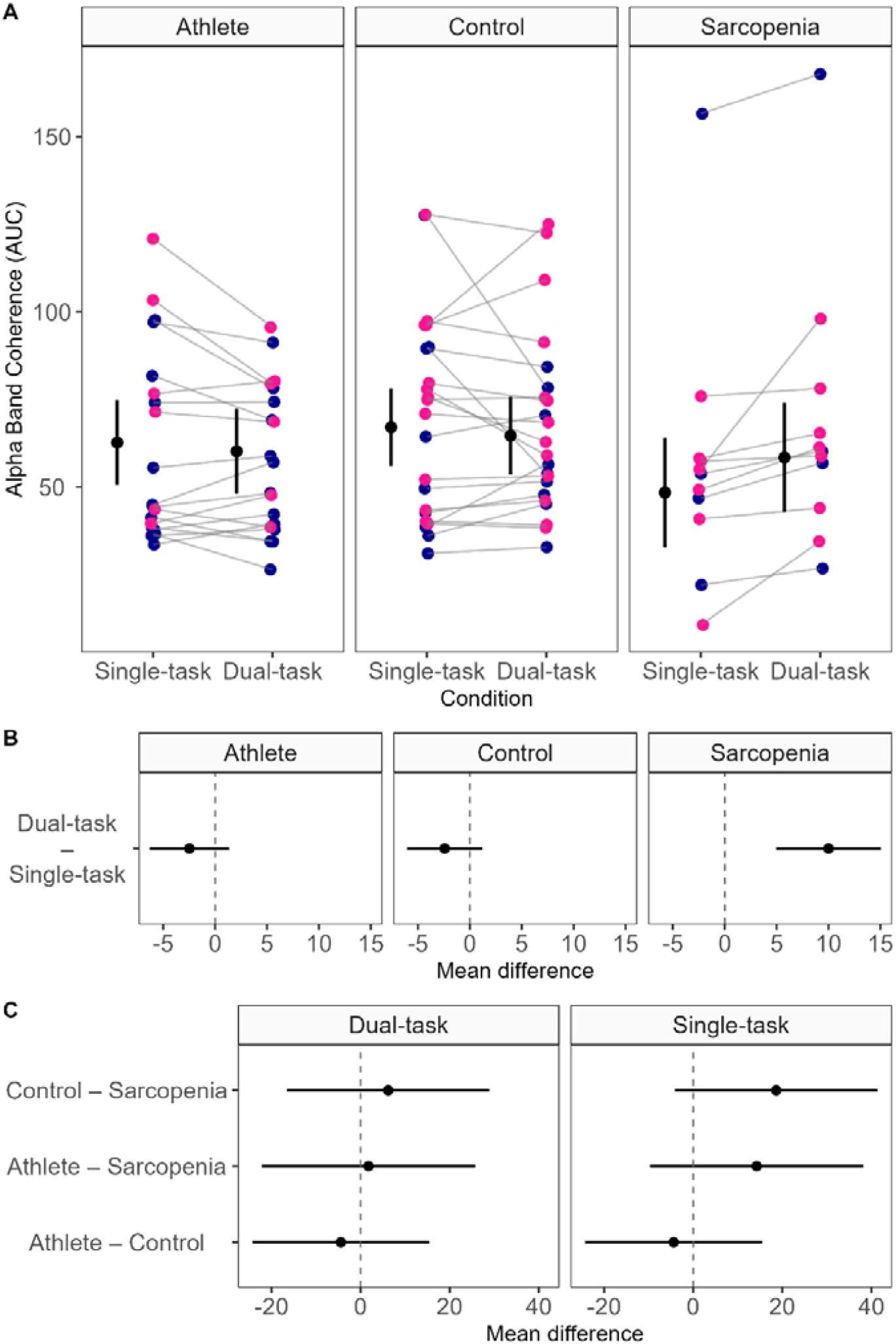
Effects of dual-tasking on alpha-band coherence across groups. Panel A displays participant-level alpha-band coherence area under the curve (AUC) during single-task and dual-task conditions for Athlete, Control, and Sarcopenia groups. Each coloured point represents the mean value for one participant (blue = male, pink = female). Panel B displays condition contrasts (dual-task – single-task) shown separately for each group. Alpha-band coherence remained unchanged in athletes and controls, but increased in sarcopenic individuals during dual-tasking. Panel C displays group contrasts, shown separately for dual- and single-task conditions. No significant differences were observed between groups in either condition. For all panels, black circles and

#### Corticospinal coupling was increased in the sarcopenic group, and reduced in controls and athletes during dual-tasking

Neural input reflecting corticospinal contributions to common drive (beta-band coherence) increased during the dual-task condition in the sarcopenic group, but decreased in controls and athletes. Figure 8 shows estimated marginal means and mean differences in beta-band coherence for each group and condition. The statistical model identified a group by condition interaction [β = -13.3 (-20.0, -6.6), t = -3.89], with no effect of sex [β = -3.7 (-15.9, 8.6), t = -0.59].

**Figure 8.**
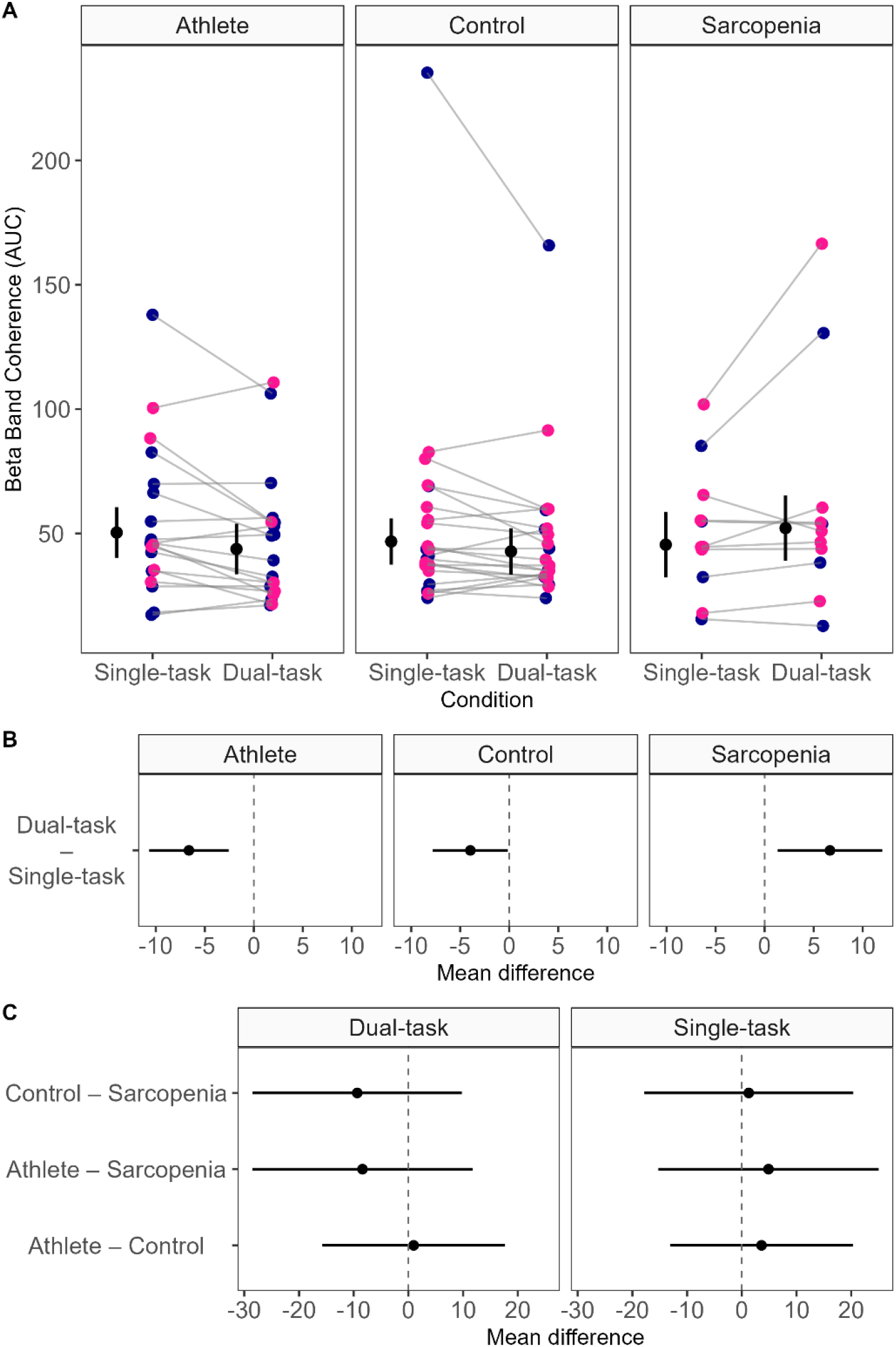
Effects of dual-tasking on beta-band coherence across groups. Panel A displays participant-level beta-band coherence area under the curve (AUC) during single-task and dual-task conditions for Athlete, Control, and Sarcopenia groups. Each coloured point represents the mean value for one participant (blue = male, pink = female). Panel B displays condition contrasts (dual-task – single-task) shown separately for each group. Beta-band coherence was significantly reduced in athletes and controls under dual-tasking, but increased in sarcopenic individuals. Panel C displays group contrasts, shown separately for dual- and single-task conditions. No significant differences were observed between groups in either condition. For all panels, black circles

### Exploratory analysis comparing endurance-vs power-type athletes

No subgroup or condition effects were found for torque coefficient of variation. Mean motor unit discharge rates increased during dual-tasking for both endurance-(n=10) and power-type (n=9) athletes. Inter-spike interval variability decreased under dual-tasking for both subgroups, although it remained consistently higher in power athletes. Regarding coherence, delta-band coherence was reduced only in power athletes during dual-tasking, whereas beta-band coherence decreased selectively in endurance athletes and was relatively higher in this subgroup under dual-tasking. Alpha-band coherence was not influenced by dual-tasking but was higher in power athletes. See Supplementary Material 2 for full results.

## DISCUSSION

This study examined whether and why force is less steady during single- and dual-task submaximal contractions in older adults with sarcopenia than non-sarcopenic controls and master athletes. Three main findings emerged. First, older adults with sarcopenia were less able to produce steady control of force, which became worse when attention was divided by a cognitive task. In contrast, controls and master athletes maintained stable performance. Second, this force unsteadiness in individuals with sarcopenia was accompanied by lower neural control steadiness, indicating that the neural signals sent to muscle also became more variable under cognitive load. Master athletes showed the opposite pattern, with better neural drive steadiness during dual-tasking, suggesting greater resilience of neuromuscular control. Third, dual-tasking changed the common synaptic input to motoneurons in different ways across groups. All groups showed lower delta-band coherence, but only the sarcopenia group showed higher alpha- and beta-band coherence, whereas controls and athletes showed lower beta-band coherence. Increased alpha-band coherence under cognitive load reflects a rise in common neural “noise” associated with physiological tremor, contributing to greater motor unit discharge variability and reduced force steadiness, indicating less efficient neural control of force production during dual-tasking. These findings suggest that impaired dual-task performance in sarcopenia is not simply a consequence of weakness but reflects reduced steadiness and altered organisation of neural drive to muscle.

### Neural control unsteadiness as a determinant of force unsteadiness in sarcopenia

Force steadiness during low-force contractions depends on the temporal regularity of motor unit discharge. Increased inter-spike interval variability has been linked to larger force fluctuations in older adults, particularly under cognitively demanding conditions^7^. In this study, participants with sarcopenia exhibited greater discharge variability during single-task contractions, which was amplified with dual-tasking, paralleling the marked deterioration in torque steadiness. Mean discharge rate increased across all groups during dual-tasking, suggesting that cognitive loading did not reduce the overall magnitude of the neural drive received by the muscles differently across groups. Rather, the key abnormality in individuals with sarcopenia appears to lie in the unsteadiness of motor unit discharge timing. This distinction is important because it suggests that the problem under cognitive stress is not simply failure to increase or sustain neural drive, but a reduced ability to organise that drive consistently enough to maintain steady force.

Dividing attentional resources is known to alter corticospinal excitability and reduce the precision of sensorimotor integration, potentially increasing variability in descending and spinal synaptic input in older adults^11,12^. In the context of sarcopenia, where motor unit remodelling, reduced motoneuron number and excitability, and impaired neuromuscular junction transmission have all been reported^4,22,38^, the motoneuron pool may operate closer to its functional limits. Including an additional cognitive demand may therefore expose instability that is partly compensated during isolated motor tasks. This may help explain why participants with sarcopenia showed a clear deterioration in force steadiness during dual-tasking whereas controls did not, despite the same general increase in mean discharge rate. Master athletes provide an informative contrast because they appear to preserve a greater capacity to maintain stable motor output when attention is divided. In our study, athletes not only preserved torque steadiness, but also reduced motor unit discharge variability during dual-tasking. Lifelong training is typically performed in dynamic environments that demand ongoing integration of sensory information, attentional shifting, decision-making, and motor execution^20,21^. Such repeated exposure may promote more efficient allocation of central resources and more adaptable control strategies^20,21^. Accordingly, the findings in this study suggest that vulnerability to motor unsteadiness under cognitive load is not an inevitable consequence of ageing itself, although differences in age, training history, and underlying health status across phenotypes should temper strong causal interpretation.

### Frequency-specific alterations in common synaptic input

Coherence analysis between motor units provided insight into how common synaptic input was modulated under cognitive load. Notably, there were no significant between-group differences in delta-, alpha-, or beta-band coherence during single-task contractions. Thus, the groups were not distinguished by large baseline differences in frequency-specific common input during the isolated motor task. Instead, the divergence emerged in how each phenotype adapted when cognitive demand was superimposed on force production. Delta-band coherence decreased in all groups during dual-tasking, suggesting that low-frequency common input was globally modulated when attentional resources are divided. Because delta-band coherence is closely related to slow force fluctuations and common drive^10,28^, this reduction may reflect a broad redistribution of neural control rather than a phenotype-specific abnormality. One possible explanation is that the addition of a cognitive demand introduces an additional source of common synaptic input to the motoneuron pool, which may reduce the relative contribution of the original low-frequency input. This interpretation is consistent with previous evidence showing that when independent common inputs are introduced to a motoneuron pool, they can decorrelate motor unit activity within specific frequency bands^39^. In contrast, alpha-band coherence showed differential dual-task responses across phenotypes. Alpha-band oscillations are associated with physiological tremor and represent components of the neural input that are not directly related to the intended force output. Because these oscillations can still be expressed in the force signal^40,41^, they contribute to unwanted fluctuations in force. Thus, they can be interpreted as a form of common “noise” in the neural drive that reduces the accuracy of force production^10^. In this study, the selective increase in alpha-band coherence in the sarcopenic group during dual-tasking provides a likely explanation for their increased motor unit discharge variability and greater torque variability and under cognitive load. Despite the reduction in delta-band coherence observed in all groups, only sarcopenic individuals showed an increase in these common “noise” oscillations, suggesting that their neural control becomes less effective at maintaining stable force when cognitive demands are increased. Beta-band coherence further highlighted differences between groups. Beta-band oscillations have been widely linked to corticospinal coupling and are thought to reflect the contribution of cortical input to the motoneuron pool^10^ Beta-band coherence increased in sarcopenic individuals during dual-tasking, which may indicate a greater reliance on cortical input when cognitive demands are increased. Conversely, the reduction in beta-band coherence observed in controls and, more prominently, in master athletes suggests a more adaptable regulation of common synaptic input. Although the present design does not permit causal inference, the emergence of these opposite modulation patterns only under dual-task conditions supports the idea that impaired neural adaptability, rather than simply impaired strength, may distinguish sarcopenic from more resilient ageing phenotypes.

### Clinical implications

The clinical importance of these findings is that sarcopenia may involve deficits in neuromuscular control that are not fully captured by measures of muscle mass or maximal strength alone. Force steadiness has been linked to mobility performance, postural control, and other clinically meaningful aspects of function^42^, and dual-task performance is strongly connected to fall risk and loss of independence in older adults^15^. In that context, the findings from this study suggest that traditional single-task strength testing may underestimate vulnerability in sarcopenic individuals when they are required to divide attention, which is precisely how many real-world activities are performed.

The findings from this study also have implications for intervention design. Resistance training is recommended to improve muscle strength in people with sarcopenia^43^, but strategies that enhance the stability and adaptability of neural drive may also be valuable. Challenging balance training, multicomponent program, dual-task training, and exercise modalities that require ongoing sensorimotor integration may be especially relevant if the goal is not only to increase force capacity, but also to improve the ability to control force under cognitively demanding conditions. In fact, dual-task exercises are known to improve dynamic balance and functional mobility, and reduces fall frequency in older adults^44^. The preserved discharge regularity and flexible modulation of common synaptic input observed in master athletes further support the concept that neural adaptability is modifiable and may represent a meaningful therapeutic target.

Force and motor unit discharge variability under cognitive load may prove useful as clinical and mechanistic markers of neuromotor vulnerability in older adults with sarcopenia. Longitudinal studies are now needed to determine whether these physiological features predict mobility decline, falls, or loss of independence, and whether interventions that improve discharge regularity also led to clinically meaningful functional outcomes. This study provides a mechanistic basis for moving beyond a purely muscle-centric view of sarcopenia toward a broader model that more fully incorporates neural control.

### Strengths and limitations

This study has several strengths. The use of HD-EMG combined with motor unit decomposition allowed high-resolution, non-invasive, characterisation of motor unit discharge behaviour and common synaptic input in vivo. Analysing discharge variability at the motor unit level provides mechanistic insight beyond traditional surface EMG amplitude measures. The inclusion of three distinct ageing phenotypes enabled examination of both vulnerability and resilience within older adults, thereby strengthening interpretation of phenotype-specific adaptations. However, several important limitations warrant consideration. First, the cross-sectional design precludes causal inference regarding whether altered discharge variability or coherence contributes directly to functional decline or instead reflects adaptation. Second, the sarcopenia group was relatively small, which limits precision and warrants caution when interpreting interaction-heavy analyses, particularly for coherence outcomes. Third, neuromuscular assessments were performed during submaximal isometric dorsiflexion of tibialis anterior; although this muscle is relevant to gait and postural control and is well suited to HD-sEMG decomposition, the findings may not generalise to other muscle groups or to more dynamic tasks. Fourth, the serial-subtraction task may not have imposed identical cognitive challenge across phenotypes and detailed cognitive task performance metrics were not recorded. Although all participants were able to perform the cognitive alone (responses faster than 3s), and task performance criteria were standardised, differential cognitive load cannot be excluded and may have influenced the magnitude of dual-task effects observed. Finally, because this study was conducted in sarcopenic individuals diagnosed using the SDOC definition^2^ the results may not generalise to individuals classified as sarcopenic under other diagnostic definitions.

## CONCLUSIONS

This study shows that older adults with sarcopenia had poorer force steadiness and greater motor unit discharge rate variability (i.e., proxy of neural control unsteadiness), and that both deficits worsened when a cognitive task was added. This pattern was not seen in healthy controls or master athletes. Mean discharge rate, however, increased similarly across all groups, suggesting that the key neural disturbance in individuals with sarcopenia under cognitive stress is not the overall level of neural drive, but rather worse neural control steadiness. Group differences in the modulation of common synaptic input further suggest that ageing phenotypes differ in how the nervous system adapts when attentional demands increase. In contrast, master athletes maintained more regular motor unit discharge and showed more flexible regulation of common synaptic input during dual tasking. These findings suggest that greater motor unit discharge variability during cognitive challenge may represent a neural feature of functional impairment in sarcopenia and support the inclusion of neuromuscular control measures, in addition to muscle mass and maximal strength, in clinical evaluation and intervention strategies.

## Data Availability

The dataset, R code, and packages information can be found at: https://github.com/orssatto/mucoh-sarcopenia.

## ACKNOWLEDGEMENTS

This study was funded by an Alfred Deakin Postdoctoral Fellowship from Deakin University to LBRO. BCC’s effort was supported in part by NIH/NIA R01AG067758. DS is supported by an Australian National Health and Medical Research Council (NHMRC) Investigator Grant (GNT1174886). LBRO declares the use of artificial intelligence for optimising and refining the R and MATLAB codes used for data and statistical analyses and plotting.

## Notes

### Competing Interest Statement

The authors have declared no competing interest.

### Author Declarations

Ethics committee/IRB of Deakin University gave ethical approval for this work.

## REFERENCES

1. Sayer AA, Cooper R, Arai H, Cawthon PM, Ntsama Essomba MJ, Fielding RA et al. Sarcopenia. Nat Rev Dis Primers 2024;10:1–16.

2. Bhasin S, Travison TG, Manini TM, Patel S, Pencina KM, Fielding RA et al. Sarcopenia Definition: The Position Statements of the Sarcopenia Definition and Outcomes Consortium. J Am Geriatr Soc 2020;68:1410–1418.

3. Kirk B, Cawthon PM, Arai H, Ávila-Funes JA, Barazzoni R, Bhasin S et al. The Conceptual Definition of Sarcopenia: Delphi Consensus from the Global Leadership Initiative in Sarcopenia (GLIS). Age Ageing 2024;53:1–10.

4. Clark B. Neural Mechanisms of Age-related Loss of Muscle Performance and Physical Function. J Gerontol A Biol Sci Med Sci 2023;78:8–13.

5. Clark BC. Neuromuscular Changes with Aging and Sarcopenia. Journal of Frailty and Aging 2019;8:7–9.

6. Manini TM, Clark BC. Dynapenia and aging: An update. Journals of Gerontology - Series A Biological Sciences and Medical Sciences 2012;67 A:28–40.

7. Hunter SK, Pereira XHM, Keenan KG. The aging neuromuscular system and motor performance. J Appl Physiol 2016;121:982–995.

8. Orssatto LB da R, Wiest MJ, Diefenthaeler F. Neural and musculotendinous mechanisms underpinning age-related force reductions. Mech Ageing Dev 2018;175:17–23.

9. Jones EJ, Chiou SY, Atherton PJ, Phillips BE, Piasecki M. Ageing and exercise-induced motor unit remodelling. J Physiol 2022;600:1839–1849.

10. Farina D, Negro F. Common Synaptic Input to Motor Neurons, Motor Unit Synchronization, and Force Control. Exerc Sport Sci Rev 2015;43:23–33.

11. Pereira HM, Schlinder-Delap B, Nielson KA, Hunter SK. Force steadiness during a cognitively challenging motor task is predicted by executive function in older adults. Front Physiol 2018;9.

12. Pereira HM, Schlinder-Delap B, Keenan KG, Negro F, Farina D, Hyngstrom AS et al. Oscillations in neural drive and age-related reductions in force steadiness with a cognitive challenge. J Appl Physiol 2019;126:1056–1065.

13. Pereira HM, Hunter SK. Cognitive challenge as a probe to expose sex- and age-related differences during static contractions. Front Physiol 2023;14.

14. Enoka RM, Farina D. Force steadiness: From motor units to voluntary actions. Physiology 2021;36:114–130.

15. Verghese J, Kuslansky G, Holtzer R, Katz M, Xue X, Buschke H et al. Walking While Talking: Effect of Task Prioritization in the Elderly. Arch Phys Med Rehabil 2007;88:50.

16. Chen H-C, Schultz AB, Ashton-Miller JA, Giordani B, Alexander NB, Guire KE. Stepping Over Obstacles: Dividing Attention Impairs Performance of Old More Than Young Adults. Journal of Gerontology: Medical Sciences 1996;51A:116–122.

17. G.E. Stelmach, H. N. Zelznik, D. Lowe. The influence of aging and attentional demands on recovery from postural instability. Aging 1990;155–191.

18. Zhao Y, Zhang J, Yang K, Liu M, Wang L, Yang C et al. The effects of cognitive-motor and motor-motor dual tasks on gait performance and dynamic stability in older adults with and without sarcopenia: a cross-sectional study. BMC Geriatr 2026;26.

19. Woollacott M, Shumway-Cook A. Attention and the control of posture and gait: a review of an emerging area of research. Gait Posture 2002;16:1–14.

20. Glenn JM, Vincenzo J, Canella CK, Binns A, Gray M. Habitual and maximal dual-task gait speeds among sedentary, recreationally active, and masters athlete late middle-aged adults. J Aging Phys Act 2015;23:433–437.

21. Dupuy O, Bosquet L, Fraser SA, Labelle V, Bherer L. Higher cardiovascular fitness level is associated to better cognitive dual-task performance in Master Athletes: Mediation by cardiac autonomic control. Brain Cogn 2018;125:127–134.

22. Orssatto LBR, Scott D, Clark BC, Lim J, Daly RM. Intrinsic Motoneuron Excitability Differentiates Sarcopenic, Nonsarcopenic and Athletic Ageing Phenotypes. J Cachexia Sarcopenia Muscle 2025;16.

23. Lecce E, Casolo A, Nuccio S, Felici F, Bazzucchi I. Analysis of motor units with high-density surface electromyography: methodological considerations and physiological significance. Eur. J. Appl. Physiol. 2025 doi:10.1007/s00421-025-05996-8.

24. Datta AK, Farmer SF, Stephens JA. Central nervous pathways underlying synchronization of human motor unit firing studied during voluntary contractions. Journal of Physiology 1991;432:401–425.

25. Miles TS, Box G. The cortical control of motor neurones: some principles of operation. Med Hypotheses 1987;23:43–50.

26. De Luca CJ, Lefever RS, Mccue MP, Xenakis AP. Control scheme governing concurrently active human motor units during voluntary contractions. J Physiol 1982;329:129–142.

27. Negro F, Holobar A, Farina D. Fluctuations in isometric muscle force can be described by one linear projection of low-frequency components of motor unit discharge rates. Journal of Physiology 2009;587:5925–5938.

28. Negro F, Farina D. Linear transmission of cortical oscillations to the neural drive to muscles is mediated by common projections to populations of motoneurons in humans. Journal of Physiology 2011;589:629–637.

29. Malmstrom TK, Miller DK, Simonsick EM, Ferrucci L, Morley JE. SARC-F: A symptom score to predict persons with sarcopenia at risk for poor functional outcomes. J Cachexia Sarcopenia Muscle 2016;7:28–36.

30. da Silva ME, Orssatto LB da R, Bezerra E de S, Silva DAS, Moura BM de, Diefenthaeler F et al. Reducing measurement errors during functional capacity tests in elders. Aging Clin Exp Res 2018;30:595–603.

31. Holobar A, Zazula D. Multichannel blind source separation using convolution Kernel compensation. IEEE Transactions on Signal Processing 2007;55:4487–4496.

32. Frančič A, Holobar A. Motor Unit Tracking Across Low Contraction Levels of Biceps Brachii Muscle. In: Torricelli D, Metin A, Pons JL, editors. Converging Clinical and Engineering Research on Neurorehabilitation IV. Springer; 2022. pp. 401–405.

33. Negro F, Muceli S, Castronovo AM, Holobar A, Farina D. Multi-channel intramuscular and surface EMG decomposition by convolutive blind source separation. J Neural Eng 2016;13.

34. Negro F, Farina D. Factors Influencing the Estimates of Correlation between Motor Unit Activities in Humans. PLoS One 2012;7:1–14.

35. Cabral H V., Cudicio A, Bonardi A, Vecchio A Del, Falciati L, Orizio C et al. Neural Filtering of Physiological Tremor Oscillations to Spinal Motor Neurons Mediates Short-Term Acquisition of a Skill Learning Task. eNeuro 2024;11.

36. Castronovo AM, Negro F, Conforto S, Farina D. The proportion of common synaptic input to motor neurons increases with an increase in net excitatory input. J Appl Physiol 2015;119:1337–1346.

37. Gallet C, Julien C. The significance threshold for coherence when using the Welch’s periodogram method: Effect of overlapping segments. Biomed Signal Process Control 2011;6:405–409.

38. Sarto F, Franchi M V., McPhee JS, Stashuk DW, Paganini M, Monti E et al. Neuromuscular impairment at different stages of human sarcopenia. J Cachexia Sarcopenia Muscle 2024;15:1797– 1810.

39. Negro F, Farina D. Decorrelation of cortical inputs and motoneuron output. J Neurophysiol 2011;106:2688–2697.

40. Bawa P, Stein RB. Frequency response of human soleus muscle. J Neurophysiol 1976.

41. Baldissera F, Cavallari P, Cerri G. Motoneuronal pre-compensation for the low-pass filter characteristics of muscle. A quantitative appraisal in cat muscle units. Journal of Physiology 1998;611–627.

42. Pereira HM, Spears VC, Schlinder-Delap B, Yoon T, Nielson KA, Hunter SK. Age and sex differences in steadiness of elbow flexor muscles with imposed cognitive demand. Eur J Appl Physiol 2015;115:1367–1379.

43. Beckwée D, Delaere A, Aelbrecht S, Baert V, Beaudart C, Bruyère O et al. Exercise interventions for the prevention and treatment of sarcopenia. A systematic umbrella review. J Nutr Health Aging 2019.

44. Khan MJ, Fong KNK, Wong TWL, Tsang WW nam, Chen C, Chan WC et al. Effectiveness of dual-task exercise in improving balance and preventing falls among older adults: systematic review with meta-analysis and meta-regression. Eur. Geriatr. Med. 2025;16:2047–2083.

